# Modelling the influence of changes in vaccination timing, timeliness and coverage on the example of measles outbreaks in the UK between 2010-19

**DOI:** 10.1101/2024.11.20.24317639

**Authors:** Anne M Suffel, Charlotte Warren-Gash, Helen I McDonald, Adam Kucharski, Alexis Robert

## Abstract

**Background:** The Measles-Mumps-Rubella vaccine is given as a two-dose course in childhood, but the schedule of the second dose varies between countries. England recommended bringing forward the second dose from three years and four months to 18 months by 2025. We aim to quantify how changing the vaccine schedule could impact measles transmission dynamics.

**Methods:** We used a mathematical model stratified by age group and region to generate stochastic outbreaks with different vaccine schedules. We used detailed information on vaccine uptake for different age groups by region and year from electronic health records and modelled alternative scenarios changing the timing of the second MMR dose or changing uptake of either dose. We simulated measles incidence between 2010 and 2019 and compared the number of cases in each scenario.

**Results and discussion:** Delivering the second MMR vaccine at younger age resulted in a lower number of cases than in the reference set of simulations with 16% (IQR: 1.93– 28.48%) cases averted when the second dose was given at 18 months. The number of cases decreased even if the coverage of the second dose decreased by up to 3% (median reduction 15.94%; IQR: 0.41 −28.21%). The impact on case numbers was equivalent to increasing first dose coverage by 0.5% every year between 2010 and 2019 (16.38 % reduction, IQR:1.90 - 28.45), more cases could be avoided (28.60%, IQR: 17.08 - 38.05) if the first dose coverage was increased by 1% every year.

Our data highlighted how patterns of vaccination uptake translate into outbreak risk. Although increasing coverage of the first MMR dose led to the best results, this may be challenging to achieve requiring substantial resources with already high coverage of the first dose. Hence, an earlier second MMR dose presents a good alternative for mitigating the risk of measles outbreaks.

## 1. Introduction

Measles is a highly contagious disease that can lead to severe illness affecting almost every organ system (1). However, measles infection is prevented very effectively by vaccination. The global eradication of measles through vaccination is feasible and has been achieved in large geographical areas (2). The first measles vaccine was developed in 1968 (3), and many countries now use the Measles-Mumps-Rubella (MMR) vaccine. A single dose of measles-containing vaccine was shown to be at least 95% effective in preventing clinical measles in preschool children. The first dose of vaccine (MMR1) should be administered around the first birthday, as earlier first dose vaccination can reduce the vaccine effectiveness (4). A second dose (MMR2) is routinely recommended as not all children respond to MMR1(3). The guidance on timing of MMR2 varies across countries and is recommended by the World Health Organization between the second year of life and school entry (3): Some European countries (e.g. France, Germany) administer MMR2 before the second year of life, while others (e.g. Finland, Latvia or Poland) only recommend MMR2 around school entry between the ages 5 to 7 years (5). Since 1996, the vaccine schedule recommended by the National Health Service in the United Kingdom states that MMR should be given at the ages of one year and at three years and four months (6,7). It remains unclear how these different vaccination strategies impact measles dynamics in low-incidence settings such as the European region.

Since measles is highly infectious, high vaccine coverage is required to mitigate the risks of outbreaks – a study by Funk et al. (2019) estimated that a coverage of 95% in five-year-olds was necessary to ensure an elimination of measles transmission (8). Uptake of the MMR vaccine in England declined in the late 90s until the early 2000s, ultimately reaching 80%, and then increased again until 2015 with additional catch-up campaigns (9). However, the coverage has been declining since 2015 (10) and was additionally impacted by the COVID-19 pandemic (11,12). These variations have been associated with several large outbreaks (above 1,000 annual cases) in the UK since 2010 (13–15).

The UK Joint Committee on Vaccination and Immunisation (JCVI) published a recommendation to bring MMR2 forward to the age of 18 months by 2025 in order to improve coverage (16). This decision was based on a study by Lacy at al indicating that an earlier vaccination schedule in some London boroughs led to 3.3% (95% CI: 1.3-5.3%) higher uptake in London at the age of five (17). However, it remains uncertain how changes in vaccination schedule would influence vaccine uptake in the long term. In particular, shifting MMR2 to the age of 18 months would require an additional vaccination appointment, which could lead to a lower acceptability or might be difficult to organise for parents which might lead to lower coverage(18).

We aimed to quantify the impact of different measles vaccination strategies on the risk of measles outbreaks in England including the existing and new vaccination schedules in the United Kingdom and alternative schedules from other European countries. (19)

## 2. Methods

### 2.1 Study design

We simulated outbreak risk under different vaccination strategies and coverage between 2010 and 2019. We chose this period to avoid the potential disruptions of the routine immunisation programme associated with the COVID-19 pandemic. We generated stochastic simulations using a mechanistic transmission model that had previously been fitted to England case data (19). We took the parameter estimates from the model fits then simulated stochastic outbreaks between 2010 and 2019. The modelled scenarios included vaccination schedules from other European countries such as introducing MMR2 at an earlier age, or recommending a later MMR2 at school entry, improving coverage of MMR1 and MMR2 separately, and changes of schedule and coverage together.

### 2.2 Outbreak data

Data on all laboratory-confirmed measles cases in England between 2010 and 2019 were obtained from Public Health England (now UK Health Security Agency). This dataset included the date of onset, region of residence, age and vaccination status of the 7504 cases and was used to fit the transmission model (for a summary see figure S1 in the supplement).

### 2.3 Vaccine data

Two data sources of vaccine coverage were used: The Clinical Practice Research Datalink (CRPD) Aurum is a primary care dataset from general practitioner (GP) surgeries practices using EMIS Web® software containing patient-level information on symptoms and diagnoses, clinical tests and results, immunisations, prescriptions and referrals to other services (20). In 2022, CPRD Aurum contained data from around 25 million patients and was broadly representative of England by geographical spread, age, sex and ethnicity (21).

Using a validated algorithm to identify vaccination records (22), vaccination coverage at the ages of 1, 2, 3, 4, and 5 years was estimated from the electronic health records. The results were previously published (23). As the CPRD data were only available for children born between 2006 and 2015, this covered all age bands between 0 and 5 in the years 2010 to 2016.

Cover of Vaccination Evaluated Rapidly (COVER) is a dataset published by NHS Digital summarising UK vaccination coverage at the age 2 and 5 for the MMR vaccine (24). At the time of the study, it was available for children born between 2000 and 2019. However, the 2-year coverage was only available for children born in 2004 and older, while the 5-year coverage was only available for children born before 2017.

For children born before 2006 or after 2015, the CPRD vaccine data had to be supplemented with estimated values based on COVER data. COVER is based on aggregated GP information based on operational data which may be incomplete, not fully representative and not quality assured (25). CPRD is closer to the corrected COVER data as it uses individual-level vaccination data. A comparison between COVER data and CPRD data can be found in the supplementary material (see figures S2, S3 and table S4 in the supplement). Based on this, COVER estimates were adjusted for 50% underascertainment (i.e. assuming that 50% of the unvaccinated children were vaccinated but did not have their vaccine recorded) (9). These corrected values were consistent with estimates from previous studies (23).

To estimate the vaccine coverage in the missing age bands, we applied the relative difference of the age bands for the last completely available years, i.e., 2006 and 2017, to the COVER estimates to supplement the values for ages three and four. All these values were stratified by region (see Figure 1).

**Figure 1.**
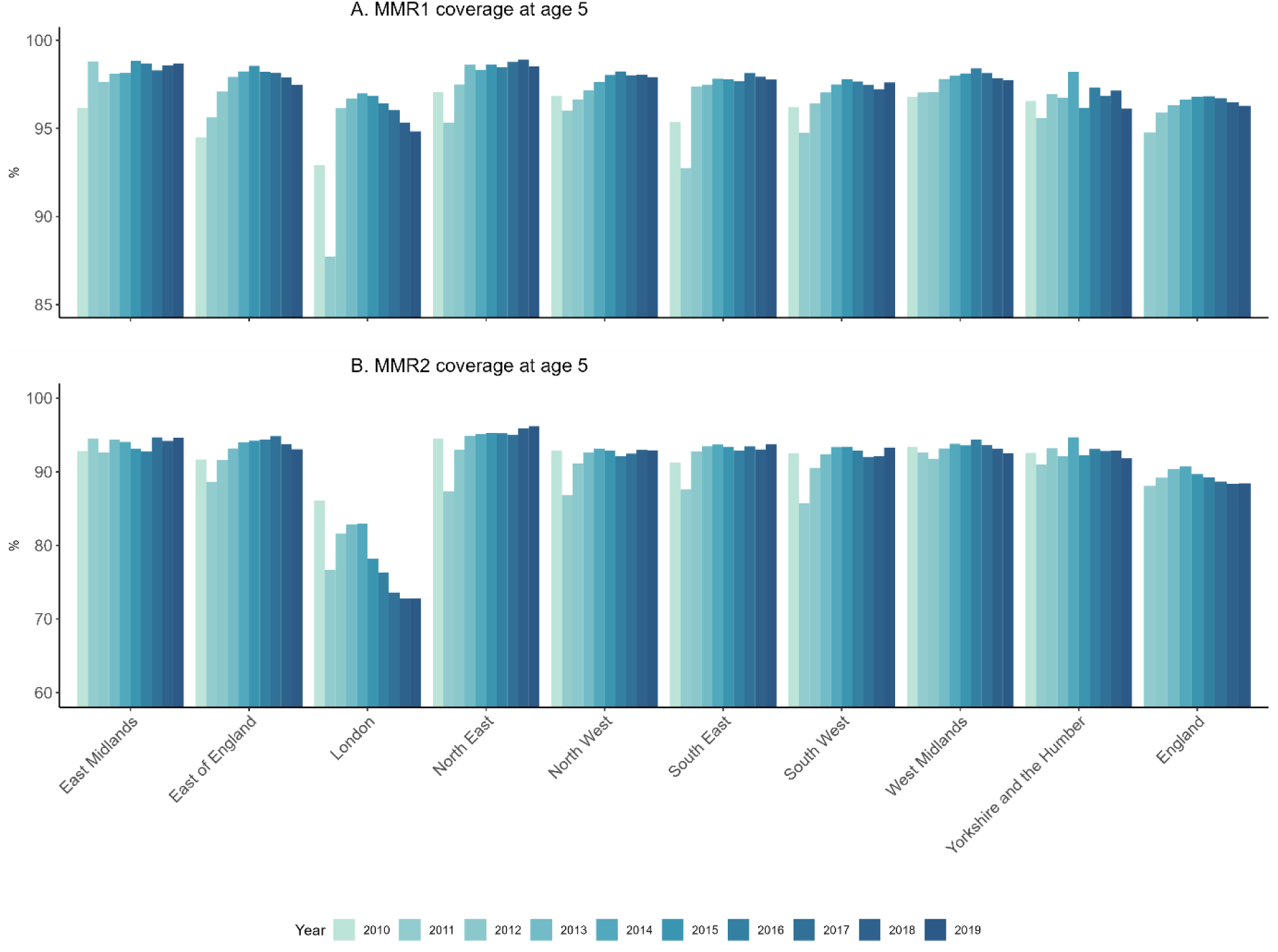
Vaccination coverage estimates stratified by region for the MMR1 (A) and MMR2 (B) at the age of five years.

### 2.4 Transmission model

We used a compartmental transmission model with compartments for susceptible, exposed, infected and recovered individuals (SEIR model) and single and double vaccinated individuals to reproduce the measles dynamics observed in England by age group, region, and vaccine status. The parameters estimated by the model were then used to generate stochastic simulations, describing the range of dynamics under a range of alternative scenarios (see Figure 2). The transmission model was described in detail in a previous publication (19)

**Figure 2.**
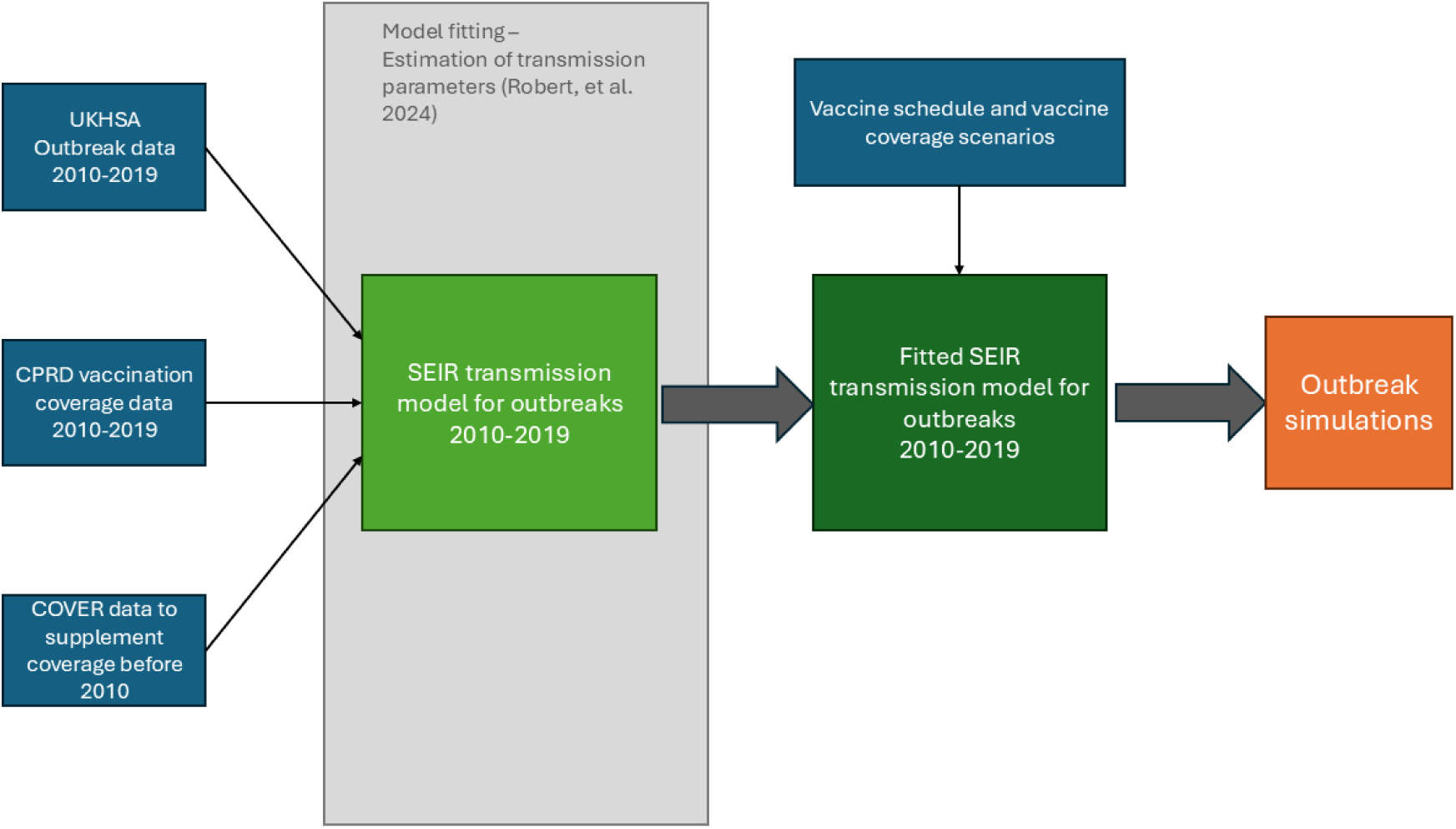
Process of fitting the compartmental transmission model and generating the outbreak simulations based on different vaccination scenarios. UKHSA: UK Health Security Agency. CPRD: Clinical Practice Research Datalink. COVER: Cover of Vaccination Evaluated Rapidly.

A more detailed description of the model fitting process and model parameters can be found in the supplementary material (see S5). We ran 250 simulations per vaccination strategy. Counterfactual scenarios were matched by using the same seed for the stochastic simulations.

### 2.5 Alternative vaccination scenarios

The vaccine data previously described were used as a reference scenario when MMR2 was given from the age of three years and four months. The scenarios were based on the current changes in the English vaccination schedule, comparing it with other European vaccination schedules, and exploring the impact of changes of coverage with and without changes in schedule. Table 1 summarises the different scenarios for which we simulated measles outbreaks in the England.

**Table 1.** Overview of the different vaccination strategies.

| Strategy name | Description of the vaccination scenario | Examples of European regions with this vaccination schedule in place (5)* |
| --- | --- | --- |
| Reference scenario | MMR1 given at one | UK schedule 2024, Cyprus, Denmark, Ireland, Slovakia |
|  | MMR2 given at three years and four months |  |
| Improving coverage of MMR1 | Same MMR1 schedule as reference<br>Same MMR2 schedule as reference<br>Increasing annual MMR1 coverage by 0.5% or 1% from age 2<br>Same MMR2 coverage as reference | UK schedule 2014 |
| Improving coverage of MMR2 | Same MMR1 schedule as reference<br>Same MMR2 schedule as reference<br>Same MMR1 coverage as reference<br>Increasing MMR2 coverage by 1 or 3% from age 4 | UK schedule 2024 |
| School Entry MMR2 | Same MMR1 schedule as reference<br>MMR2 given at school entry (5 years)<br>Same MMR1 coverage as reference<br>Same MMR2 coverage as reference | Belgium, Croatia, Finland, Latvia |
| Early MMR2 | Same MMR1 schedule as reference<br>MMR2 given at the age of two years.<br>Same MMR1 coverage as reference<br>Different sub-scenarios were included with changes in MMR2 coverage. | France, Germany, UK schedule from 2025 |

\*Some countries have wider ranges of recommended timings for the vaccine and this table does not reflect the exact timing in these countries.

We firstly created scenarios which only included an increase in coverage for MMR1 or MMR2 alone, and then looked at alternative vaccination schedules under different assumptions about how this would influence vaccine coverage. We used the median number of cases between 2010 and 2019 and the IQR in each simulation set to estimate the impact of changes in vaccination schedule and coverage. The number of cases is compared to the reference set of simulation by computing the median and IQR of the percentage of change (increase of decrease) between each simulation set and the median number of cases in the reference simulations.

#### Improving vaccine coverage

We firstly created scenarios in which the timing of MMR2 was not changed from the original schedule but the overall coverage of MMR1 or MMR2 was increased across all regions and years between 2010 and 2019. We explored an increase by 0.5 and 1% for either MMR1 and or MMR2. As coverage for MMR2 at the age of five is lower than for MMR1, we further included a scenario with increasing MMR2 by 3% or reducing coverage by up to 3%.

#### Introducing an earlier MMR2

We explored two alternative scenarios with MMR2 recommended at the age of two instead of three years and four months. In the new schedule recommended by the JCVI, MMR2 would be delivered at 18 months, but since our model is stratified by age groups (1 year-age bands until 6), we used 24 months in the simulations. The first assumed the uptake would follow the same pattern of timeliness as the current MMR2 delivery, therefore one year and four months were subtracted from all the dates when MMR2 was received. New coverage estimates were calculated from these updated dates. In the second, we assumed that MMR2 recommended at the age of two would be taken up with the same speed as MMR1, which is usually faster than MMR2 (23).

We also explored the impact of moving MMR2 recommendation to five years, assuming that the speed of uptake was similar to the reference MMR2 speed. New coverage estimates were calculated from the updated dates.

Finally, we looked at potential scenarios in which an earlier MMR2 would also influence coverage for MMR2. For this we explored an increase by 0.25, 0.5 and 1% and added a scenario in the coverage for MMR1 and MMR2 were the equal. Lastly, we added one scenario in which the coverage of MMR2 would be negatively impacted by bringing MMR2 forward in the schedule.

### 2.6 Sensitivity analysis

We conducted two different sensitivity analyses: 1) we recreated the scenarios with changes in the vaccination schedule using uncorrected COVER data instead of CPRD data, and 2) we included waning of immunity from the age 5 for the scenarios with changes in schedule using the original CPRD data.

## 3. Results

In the reference scenario, our model replicated the overall pattern of measles outbreaks in England between 2010 and 2019, capturing the peak of cases in 2012 and a second smaller peak in 2018 (see Figure 3). Predicted measles cases in a majority of simulations in 2013, 2016 and 2018 were below the observed outbreak data, and above the observed cases in 2014 and 2015.

**Figure 3.**
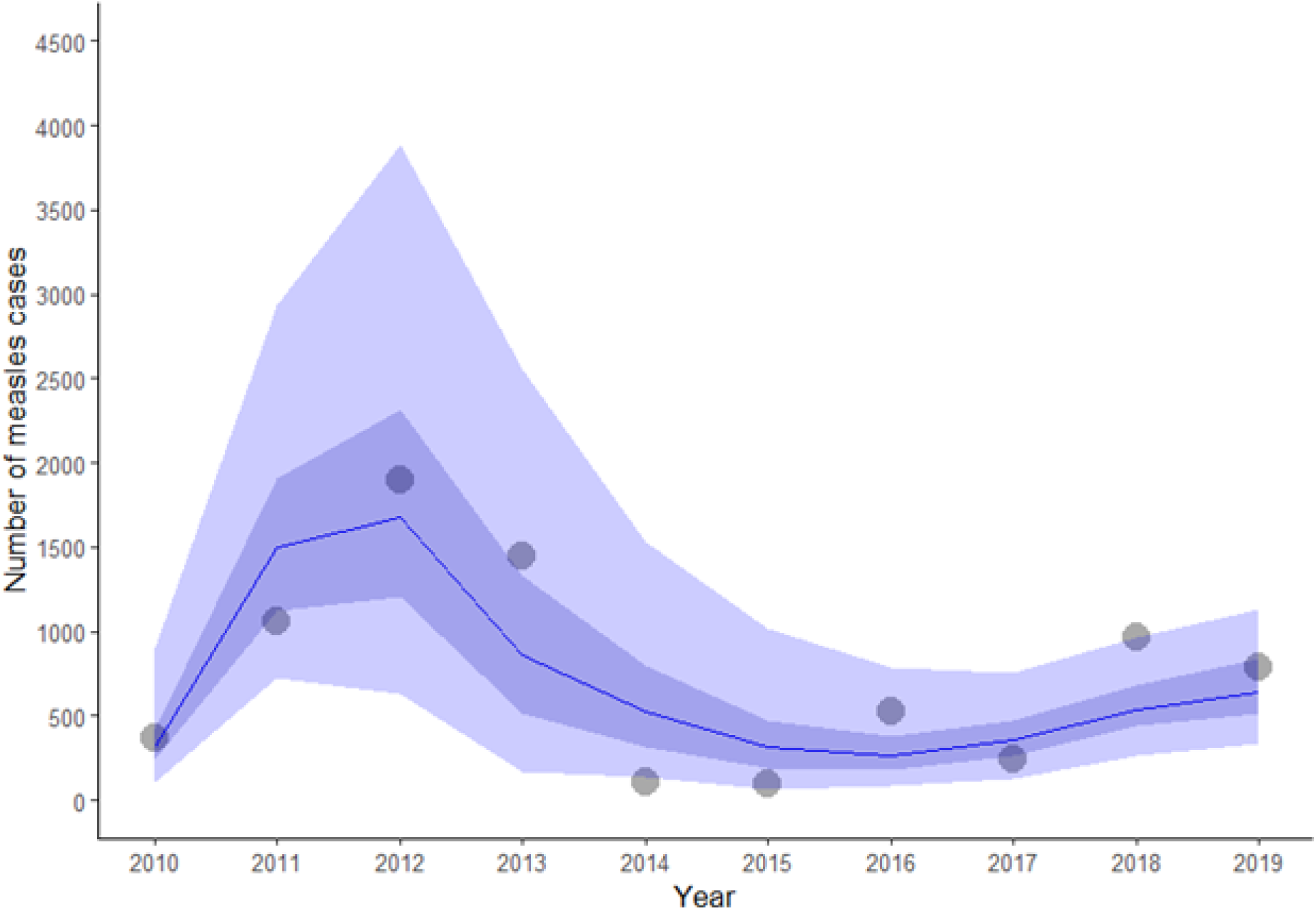
Number of notified measles cases in England (dots) and median number, interquartile range (light blue shade) and 95% simulation interval (dark blue shade) of simulated measles cases by the transmission model (blue line).

In the reference scenario, the transmission model simulated across 250 simulations showed a median of 7,081 cases (IQR: 6,008 – 8,388, see table S.7 in the supplement) between 2010 and 2019 (7502 were reported in the data).

### Changes in coverage

Improving the annual MMR1 coverage had a strong impact on reducing measles cases if achieved on sufficient scale. There was no clear difference in cases for a 0.25% improvement of coverage for MMR1 (7.88% median difference, IQR: −8.79 −21.98 %), a 0.5% improvement led to a 16.38 % reduction of cases and an improvement by 1% led to 28.60 % averted cases (see Table S.7 and Figure 4). Improvements of MMR2 coverage of up to 3% by year had no effect on avoiding measles cases (see Figure 4).

**Figure 4.**
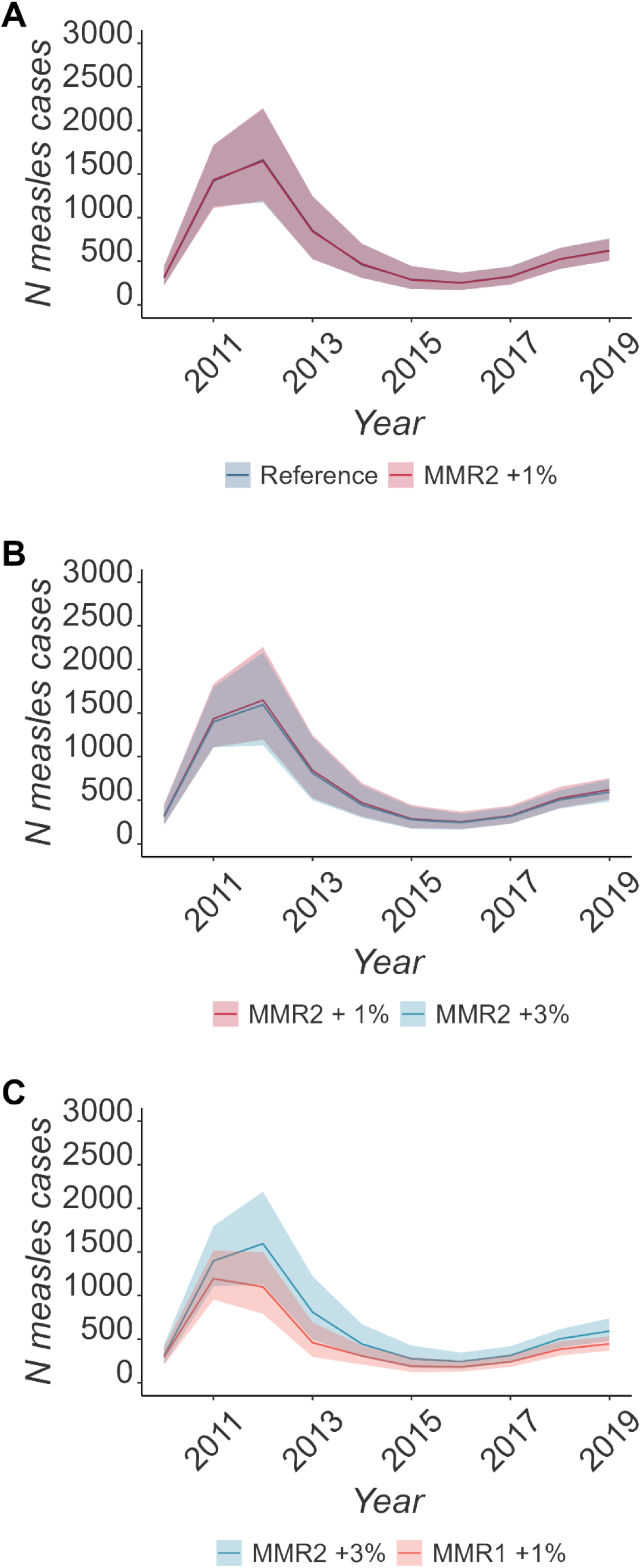
Comparing the median number and IQR of simulated cases across simulations between (A) Reference scenario and MM2 given at school age, (B) Reference scenario and MMR2 given at the age of two, (C) MMR2 given at the age two against an increase of MMR1 by 1%.

### Changes in timing

Moving MMR2 to two years instead of three years and four months reduced retrospectively the overall number of cases by 16.00% (IQR: 1.93– 28.48%) in comparison to the median number of cases in the reference scenarios (see Figure 5). If the speed of roll out of the earlier MMR2 was similar to MMR1, up to 16.54% (IQR: 2.14 – 28.69%) of cases were adverted.

**Figure 5.**
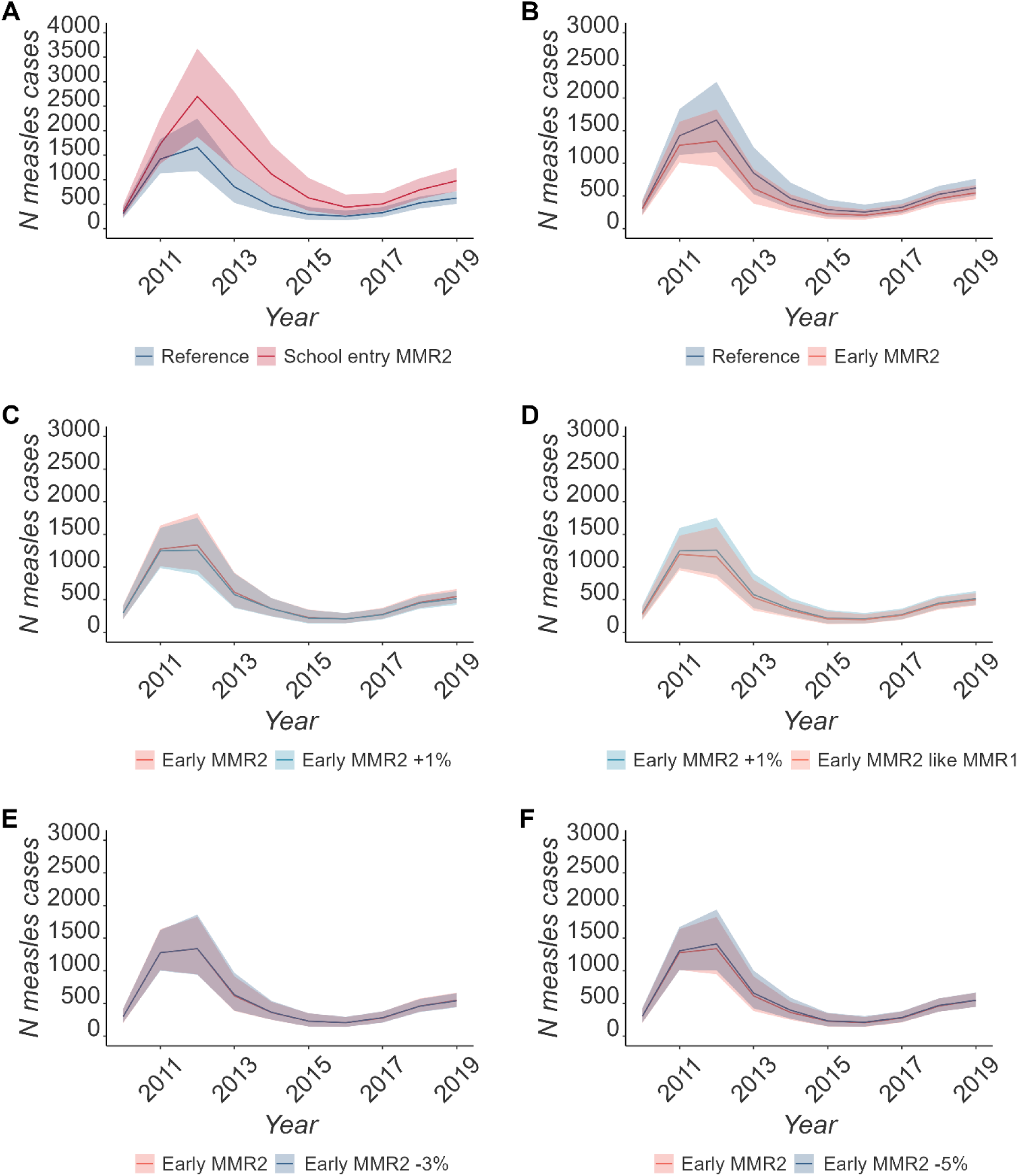
Comparing the median number and IQR of simulated cases across simulations using CPRD data and including waning between (A) Reference scenario and MM2 given at school age, (B) Reference scenario and MMR2 given at the age of two, (C) MMR2 given at the age two against an increase of MMR1 by 1%., (D) increased MMR1 by 1% and an earlier MMR2 with the same coverage as MMR1. (E) and (F) are comparing the early MMR2 with the same uptake as before against a drop in coverage by 3% and 5% respectively.

Moving MMR2 to school entry at the age of 5 increased the number of simulated measles cases by 67.61 % (IQR: −106.87 - −34.74%). Bringing forward MMR2 gave comparable estimates of cases averted, regardless of vaccine coverage (see table S.7 in the supplement, figure 4 E-F)

### Comparing timing and coverage

Both increasing the coverage of MMR1 by more than 0.25% and bringing MMR2 forward led to a reduction in measles cases across the simulations. An earlier MMR2 achieved comparable results to increasing the coverage of MMR1 to 0.5% but performed slightly worse than improving MMR1 by 1% (see table S7). These findings were relatively robust to changes in coverage for MMR2 in combination with an earlier administration (see figure 4 C-F). Delaying MMR2 at the school entry age of 5, performed worse than all the other vaccination schedule scenarios.

### Changes in age structure of measles cases

The different vaccination strategies had an influence on the age distribution of measles cases.

MMR2 given at school age increased measles cases across all age groups and almost doubled the number of cases in children between 4-6 years in comparison to the reference scenario (see Figure 6.C and F). An earlier MMR2 led to a lower proportion of measles cases in the children between 2-4 years. An improved uptake for MMR1 by 0.5% which showed a similar median reduction of cases as the early MMR2 scenario decreased the number of measles cases across all age groups (see Figure 6).

**Figure 6.**
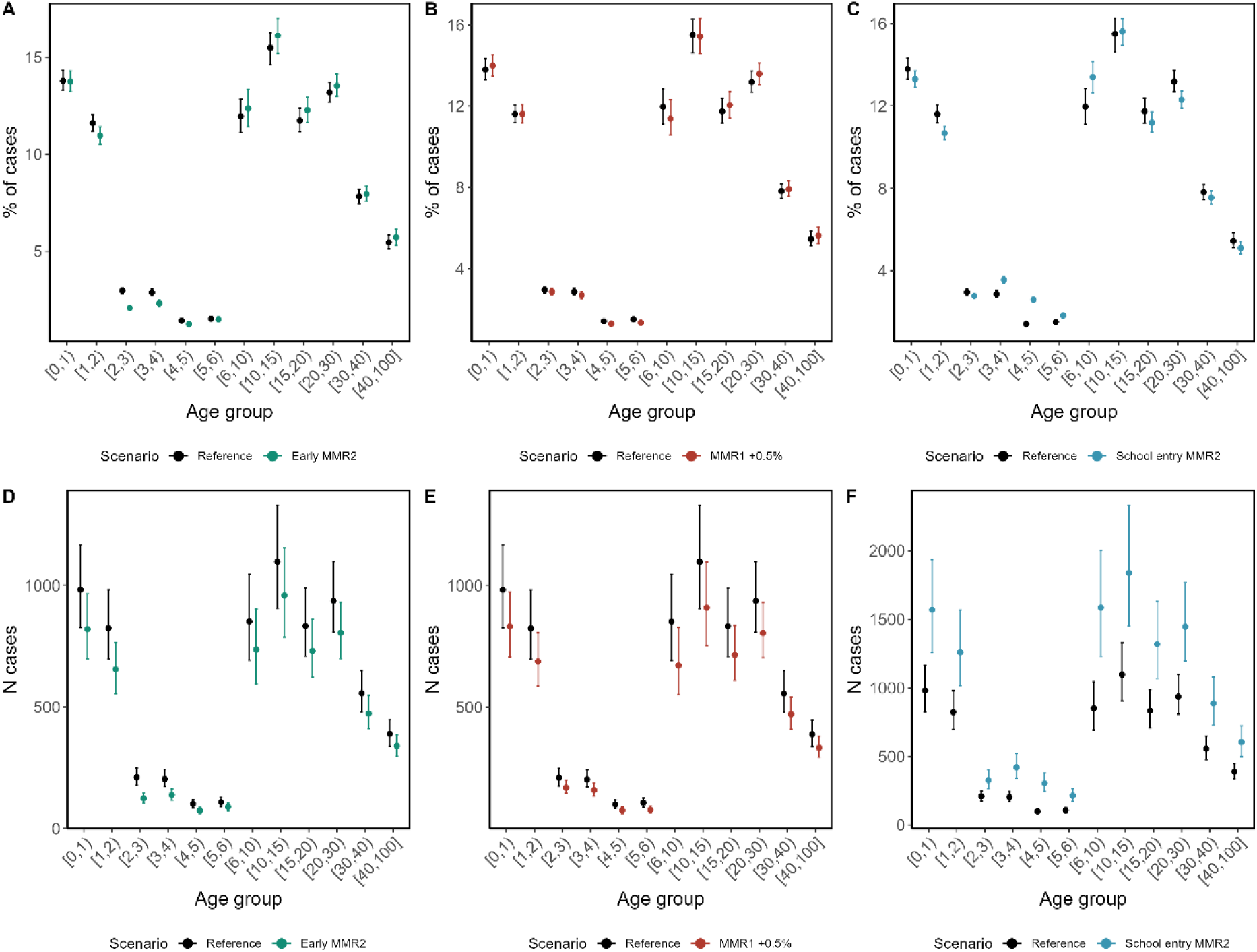
Comparison of measles cases per age compartment as proportion of all cases and absolute numbers of cases between the reference scenario and an earlier MMR2 (A+D), between reference scenario and MMR1 improved by 0.5% (B+E) and MMR2 given at the age of five (C+F).

### Sensitivity analysis

When using COVER data instead of CPRD data, there was a lower reduction in cases with a 9.66% (IQR: −7.03; 23.78%) difference in cases when MMR2 was given at age 2 in comparison to the median of the reference scenario (see Figure S8 and Table S9). Including waning to the model led to a similar median reduction of cases for the same comparison with more uncertainty (14.45% reduction, IQR: −1.48; 26.52%; see Figure S10 and Table S11).

## Discussion

We explored how different vaccination schedules could impact the risk and dynamics of measles outbreaks and showed that bringing MMR2 forward could lead to a reduction in case numbers comparable to increasing MMR1 coverage by 0.5%. Both resulted in a16% reduction of cases (IQR for 0.5% increase MMR1: 1.90 – 28.45%; IQR for early MMR2: 1.93 – 28.48%) between 2010 and 2019. These changes in the vaccine schedule could lead to significant short-term improvement and may mitigate risks of large outbreaks. This decrease was observed even if changing the vaccine schedule led to decreasing MMR2 coverage by up to 3%.

Immediate interventions to improve uptake such as the catch-up campaigns in London led to a significant improvement in coverage at the time (17), though the coverage decreased again in longer term (23,24). Bringing MMR2 forward as part of the regular NHS immunisations schedule could be a more sustainable intervention than short-term campaigns. However, this only works under the assumption that an additional vaccination appointment will be accepted by parents as the MMR2 is currently given at the same time as the 4-in-1 preschool booster. Determinants of childhood vaccine uptake are complex, and include parental vaccine confidence, opportunities to ask questions, reminders and barriers to vaccination appointments, which could all be impacted by changes in vaccine schedules (26). However, vaccine uptake is generally higher for the earlier appointments in the UK childhood vaccination schedule and so it may be reasonable to assume that an earlier MMR2 date is likely to result in higher uptake (23)

Improving the MMR2 coverage alone did not influence the outbreak dynamics. This suggests that it is most important to focus on unvaccinated individuals where possible to mitigate the risk of outbreaks.

Although moving MMR2 to the age of five resulted in more cases than other scenarios, there are pragmatic factors which might make this option favourable: The administration of a vaccine in a school setting is convenient for families and offers peer-support and opportunities for education related to personal health and health care (27,28). However, we previously observed lower vaccine uptake for appointments scheduled later in the routine immunisation schedule (23)

Strengths of this study included the use of a compartmental transmission model estimating the intensity of transmission. This approach allowed us to test different hypothetical scenarios in detail which would not be possible in real life due to financial and logistical constraints. We also used detailed, representative data on vaccination uptake which allowed to study changes in uptake by age more precisely.

There are several limitations to this approach. Firstly, the model lacks spatial granularity. Outbreaks in low incidence settings are usually driven by heterogeneity in vaccination in certain groups and communities that cannot be captured on a regional scale (29,30). However, to protect anonymity the vaccine data was not available on a more granular scale. Additionally, such data would be not adapted to a compartmental model as this would lead to an increase of compartments. Secondly, we assumed an immediate implementation and acceptance of the altered vaccination schedules. However, it is likely that there would a transition period from one schedule to another, hence, our model was overestimating the number of cases avoided at the start of the simulation.

The period of the simulation covered only ten years and does not account for long-term effects of changing the age of vaccine receipt. This might be especially important as recent evidence suggested a potential waning of immunity from the MMR vaccine (31,32). Hence, a closer monitoring of long-term immunity for measles in countries of near-elimination settings is needed. Finally, a second vaccine dose given at younger age might lead to marginally less protection in adults in the long term (19).

For policy makers, this work provided insight into different trade-offs when making decisions related to vaccination. While we found that increasing MMR1 coverage would be most effective at reducing cases, this may be challenging as interventions to improve vaccine uptake are usually very context-specific and have to be tailored to vulnerable groups in the population with mixed evidence for general interventions such as easier access and reminders (33). The COVID-19 pandemic increased vaccine hesitancy in parents towards general childhood immunisations (34) which presents an additional challenge for improving coverage. Bringing the second dose forward to the age of two or younger could improve uptake at younger age as childhood immunisations show better uptake the earlier that they are recommended in the immunisation schedule (23). With the appointment at three years and four months still in place for the preschool booster, this would still provide an additional opportunity of health care contact with children and their parents to catch up on an MMR dose which may have been missed before.

## Authors contribution

AS, CWG, HM, AJK and AR developed the analysis plan. AS and AR implemented the analysis, wrote the code, and ran the model. AS computed and collated the coverage data. AS and AR interpreted the results. AS wrote the first draft and the additional file. AS, AR, CWH, HM and AJK contributed to the manuscript and read and approved the final version of the manuscript

## Data availability statement

The study uses data from the Clinical Practice Research Datalink (CPRD). CPRD does not allow the sharing of patient-level data. The data specification for the CPRD data set is available at: https://cprd.com/cprd-aurum-may-2022-dataset. The COVER data is publicly available: https://www.england.nhs.uk/statistics/statistical-work-areas/child-immunisation/.

The analysis code can be found in the following GitHub repository: https://github.com/alxsrobert/measles_england_sir/tree/vaccination_scenarios/R

## Conflicts of Interest Statement

There were no conflicts of interest to declare.

## Acknowledgements

This work uses data provided by patients and collected by the NHS as part of their care and support (usemydata.org).

## Ethics statement

We received data governance approval from CPRD (protocol number 22_001706) and ethical approval from the London School of Hygiene and Tropical Medicine’s research ethics committee (reference number 27651).

## Funding Statement

AS and HM are funded by the National Institute for Health and Care Research (NIHR) Health Protection Research Unit in Vaccines and Immunisation (NIHR200929), a partnership between UK Health Security Agency and the London School of Hygiene and Tropical Medicine. The views expressed are those of the author(s) and not necessarily those of the NIHR, UK Health Security Agency or the Department of Health and Social Care. CWG is supported by a Wellcome Career Development Award (225868/Z/22/Z). AR was supported by the National Institute for Health Research (NIHR200908), AJK was supported by a Sir Henry Dale Fellowship jointly funded by the Wellcome Trust and the Royal Society (206250/Z/17/Z).

## Notes

### Competing Interest Statement

The authors have declared no competing interest.

### Author Declarations

We received data governance approval from Clinical Practice Research Link (protocol number 22_001706) and ethical approval from the London School of Hygiene and Tropical Medicine research ethics committee (reference number 27651).

## References

1. Perry RT, Halsey NA. The Clinical Significance of Measles: A Review. Orenstein WA, editor. J Infect Dis [Internet]. 2004 May 1;189(Supplement_1):S4–16. Available from: https://academic.oup.com/jid/article/189/Supplement_1/S4/823958

2. Moss WJ, Strebel P. Biological Feasibility of Measles Eradication. J Infect Dis. 2011 Jul;204(suppl_1):S47–53.

3. World Health Organization. Measles vaccine : WHO position paper. Weekly epidemiological report [Internet]. 2017; Available from: https://iris.who.int/bitstream/handle/10665/255149/WER9217.pdf?sequence=1

4. Demicheli V, Jefferson T, Rivetti A, Price D. Vaccines for measles, mumps and rubella in children. Cochrane Database Syst Rev [Internet]. 2005;(4):CD004407. Available from: http://ovidsp.ovid.com/ovidweb.cgi?T=JS&PAGE=reference&D=med6&NEWS=N&AN=16235361

5. European Centre for Disease Control. Measles: Recommended vaccinations [Internet]. 2024 [cited 2024 May 22]. Available from: https://vaccine-schedule.ecdc.europa.eu/Scheduler/ByDisease?SelectedDiseaseId=8&SelectedCountryIdByDisease=-1#:∼:text=MMR%20vaccination%20possible%20from%209,first%20vaccination%20before%2012%20months

6. UK Health Security Agency. 21 Measles. In: The Green Book [Internet]. 2019. Available from: https://www.gov.uk/government/publications/measles-the-green-book-chapter-21

7. Department of Health, NHS England. NHS public health functions agreement 2025-16. 2014 [cited 2024 Jul 9]. Measles, mumps, and rubella (MMR) immunisation programme. Available from: https://assets.publishing.service.gov.uk/media/5a7db6ec40f0b65d88633f79/1516_No10_Measles_Mumps_and_RubellaMMRImmunisation_Programme_FINAL.pdf

8. Funk S, Knapp JK, Lebo E, Reef SE, Dabbagh AJ, Kretsinger K, et al. Combining serological and contact data to derive target immunity levels for achieving and maintaining measles elimination. BMC Med [Internet]. 2019 Dec 25;17(1):180. Available from: https://bmcmedicine.biomedcentral.com/articles/10.1186/s12916-019-1413-7

9. UK Health Security Agency. Measles: risk assessment for resurgence in the UK [Internet]. 2023. Available from: https://www.gov.uk/government/publications/measles-risk-assessment-for-resurgence-in-the-uk

10. NHS Digital & UK Health Security Agency. Childhood Vaccination Coverage Statistics - 2020-21 [Internet]. 2021 [cited 2021 Oct 12]. Available from: https://digital.nhs.uk/data-and-information/publications/statistical/nhs-immunisation-statistics/england---2020-21

11. McDonald HI, Tessier E, White JM, Woodruff M, Knowles C, Bates C, et al. Early impact of the coronavirus disease (COVID-19) pandemic and physical distancing measures on routine childhood vaccinations in England, January to April 2020. Eurosurveillance [Internet]. 2020 May 14;25(19). Available from: https://www.eurosurveillance.org/content/10.2807/1560-7917.ES.2020.25.19.2000848

12. McQuaid F, Mulholland R, Sangpang Rai Y, Agrawal U, Bedford H, Cameron JC, et al. Uptake of infant and preschool immunisations in Scotland and England during the COVID-19 pandemic: An observational study of routinely collected data. Persson LÅ, editor. PLoS Med [Internet]. 2022 Feb 22;19(2):e1003916. Available from: https://dx.plos.org/10.1371/journal.pmed.1003916

13. Ghebrehewet S, Thorrington D, Farmer S, Kearney J, Blissett D, McLeod H, et al. The economic cost of measles: Healthcare, public health and societal costs of the 2012–13 outbreak in Merseyside, UK. Vaccine [Internet]. 2016 Apr;34(15):1823–31. Available from: https://linkinghub.elsevier.com/retrieve/pii/S0264410X16001717

14. Currie J, Davies L, McCarthy J, Perry M, Moore C, Cottrell S, et al. Measles outbreak linked to European B3 outbreaks, Wales, United Kingdom, 2017. Eurosurveillance [Internet]. 2017 Oct 19;22(42). Available from: https://www.eurosurveillance.org/content/10.2807/1560-7917.ES.2017.22.42.17-00673

15. Pegorie M, Shankar K, Welfare WS, Wilson RW, Khiroya C, Munslow G, et al. Measles outbreak in Greater Manchester, England, October 2012 to September 2013: epidemiology and control. Euro Surveill [Internet]. 2014;19(49). Available from: http://ovidsp.ovid.com/ovidweb.cgi?T=JS&PAGE=reference&D=med11&NEWS=N&AN=25523970

16. Department of Health and Social Care (DHSC). Joint Committee on Vaccination and Immunisatin (JCVI) interim statement on the immunisation schedule for children [Internet]. 2022 [cited 2023 Nov 23]. Available from: https://www.gov.uk/government/publications/jcvi-interim-statement-on-changes-to-the-childhood-immunisation-schedule/joint-committee-on-vaccination-and-immunisation-jcvi-interim-statement-on-the-immunisation-schedule-for-children

17. Lacy J, Tessier E, Andrews N, White J, Ramsay M, Edelstein M. Impact of an accelerated measles-mumps-rubella (MMR) vaccine schedule on vaccine coverage: An ecological study among London children, 2012–2018. Vaccine [Internet]. 2022 Jan;40(3):444–9. Available from: https://linkinghub.elsevier.com/retrieve/pii/S0264410X21016194

18. Smith LE, Amlôt R, Weinman J, Yiend J, Rubin GJ. A systematic review of factors affecting vaccine uptake in young children. Vaccine [Internet]. 2017;35(45):6059–69. Available from: 10.1016/j.vaccine.2017.09.046

19. Robert A, Suffel AM, Kucharski AJ. Long-term waning of vaccine-induced immunity to measles in England: a mathematical modelling study. Lancet Public Health. 2024 Sep;

20. Medicines & Healthcare products Regulatory Agency. CPRD Aurum March 2021 dataset [Internet]. 2021 [cited 2021 Oct 12]. Available from: https://www.cprd.com/cprd-aurum-march-2021

21. Wolf A, Dedman D, Campbell J, Booth H, Lunn D, Chapman J, et al. Data resource profile: Clinical Practice Research Datalink (CPRD) Aurum. Int J Epidemiol [Internet]. 2019 Dec 1;48(6):1740–1740g. Available from: https://academic.oup.com/ije/article/48/6/1740/5374844

22. Suffel AM, Walker JL, Campbell CNJ, Carreira H, Warren-Gash C, McDonald HI. Methodological challenges and recommendations for identifying childhood immunisations using routine electronic health records in the United Kingdom. medRxiv. 2023;2002–23.

23. Suffel AM, Walker JL, Williamson E, McDonald HI, Warren-Gash C. Timeliness of childhood vaccination in England: A population-based cohort study. Vaccine [Internet]. 2023 Sep;41(39):5775–81. Available from: https://linkinghub.elsevier.com/retrieve/pii/S0264410X2300926X

24. NHS digital. Cover of Vaccination Evaluated Rapidly (COVER) [Internet]. 2023. Available from: https://digital.nhs.uk/data-and-information/data-collections-and-data-sets/data-collections/cover-of-vaccination-evaluated-rapidly

25. NHS digital. Cover of Vaccination Evaluated Rapidly (COVER) [Internet]. 2023 [cited 2024 Jul 25]. Available from: https://digital.nhs.uk/data-and-information/data-collections-and-data-sets/data-collections/cover-of-vaccination-evaluated-rapidly

26. Skirrow H, Lewis C, Haque H, Choudary-Salter L, Foley K, Whittaker E, et al. ‘Why did nobody ask us?’: A mixed-methods co-produced study in the United Kingdom exploring why some children are unvaccinated or vaccinated late. Vaccine. 2024 Sep;42(22):126172.

27. Lind C, Russell ML, MacDonald J, Collins R, Frank CJ, Davis AE. School-Based Influenza Vaccination: Parents’ Perspectives. PLoS One. 2014 Mar 31;9(3):e93490.

28. Paterson P, Schulz W, Utley M, Larson H. Parents’ Experience and Views of Vaccinating Their Child against Influenza at Primary School and at the General Practice. Int J Environ Res Public Health. 2018 Mar 28;15(4):622.

29. Glasser JW, Feng Z, Omer SB, Smith PJ, Rodewald LE. The effect of heterogeneity in uptake of the measles, mumps, and rubella vaccine on the potential for outbreaks of measles: a modelling study. Lancet Infect Dis [Internet]. 2016 May;16(5):599–605. Available from: https://linkinghub.elsevier.com/retrieve/pii/S1473309916000049

30. Keenan A, Ghebrehewet S, Vivancos R, Seddon D, MacPherson P, Hungerford D. Measles outbreaks in the UK, is it when and where, rather than if? A database cohort study of childhood population susceptibility in Liverpool, UK. BMJ Open [Internet]. 2017 Mar 30;7(3):e014106. Available from: https://bmjopen.bmj.com/lookup/doi/10.1136/bmjopen-2016-014106

31. Yang L, Grenfell BT, Mina MJ. Waning immunity and re-emergence of measles and mumps in the vaccine era. Curr Opin Virol [Internet]. 2020 Feb;40:48–54. Available from: https://linkinghub.elsevier.com/retrieve/pii/S1879625720300304

32. Robert A, Suffel AM, Kucharski AJ. Long-term waning of vaccine-induced immunity to measles in England. medRxiv [Internet]. 2024 [cited 2024 May 22]; Available from: https://www.medrxiv.org/content/10.1101/2024.04.18.24306028v1

33. Crocker-Buque T, Edelstein M, Mounier-Jack S. Interventions to reduce inequalities in vaccine uptake in children and adolescents aged <19 years: a systematic review. J Epidemiol Community Health (1978) [Internet]. 2017 Jan;71(1):87–97. Available from: https://jech.bmj.com/lookup/doi/10.1136/jech-2016-207572

34. He K, Mack WJ, Neely M, Lewis L, Anand V. Parental Perspectives on Immunizations: Impact of the COVID-19 Pandemic on Childhood Vaccine Hesitancy. J Community Health [Internet]. 2022 Feb 23;47(1):39–52. Available from: https://link.springer.com/10.1007/s10900-021-01017-9

