## Supplementary material for "Modelling the influence of changes in vaccination timing, timeliness and coverage on the example of measles outbreaks in the UK between 2010-19"

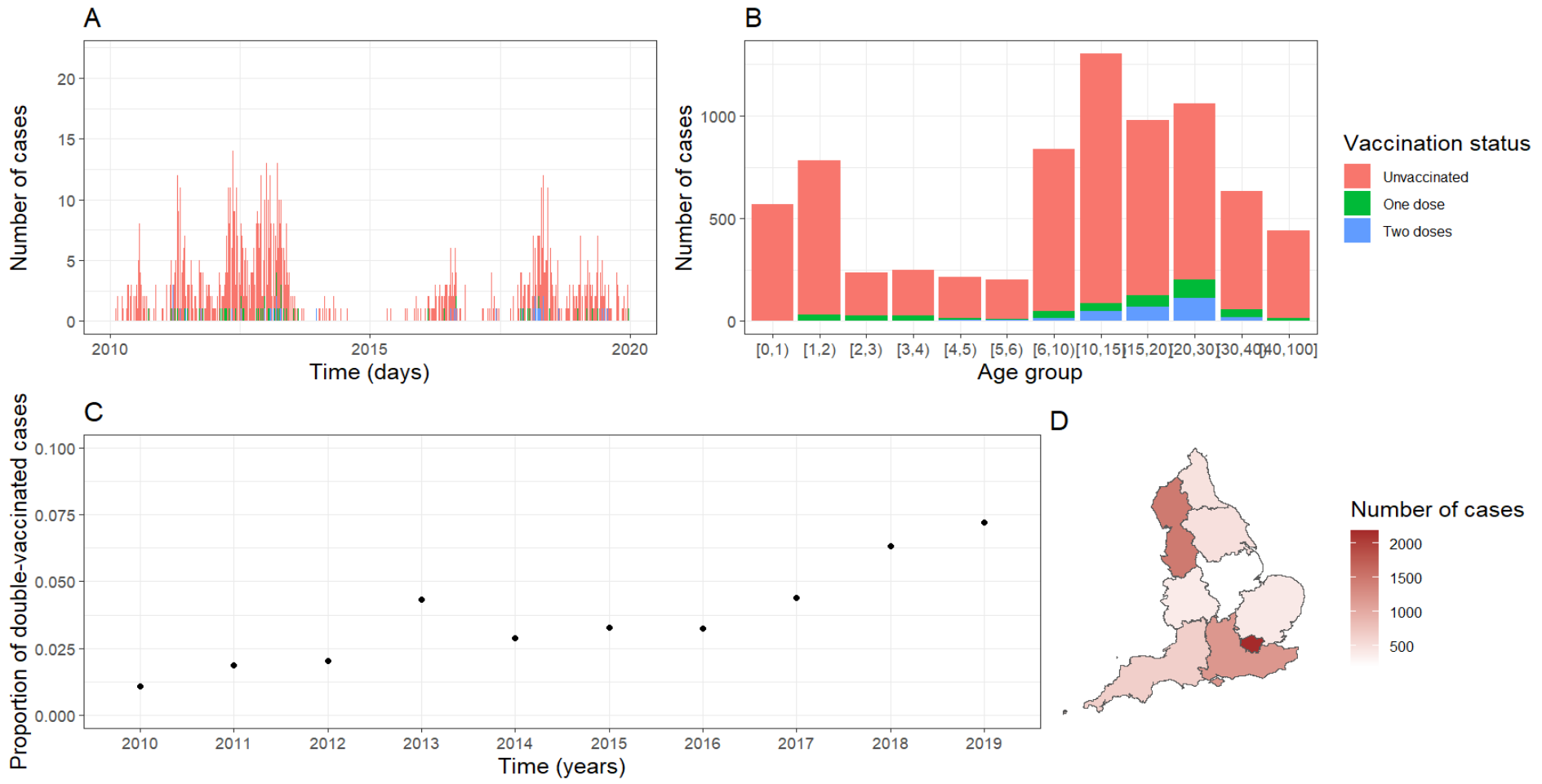

**Figure S1**. A) Number of daily measles cases in England, stratified by vaccination status. B) Number of cases by age group and vaccine status. C) Proportion of double vaccinated cases each year. D) Incidence by region of England between 2010 and 2019. Reproduced from (19)

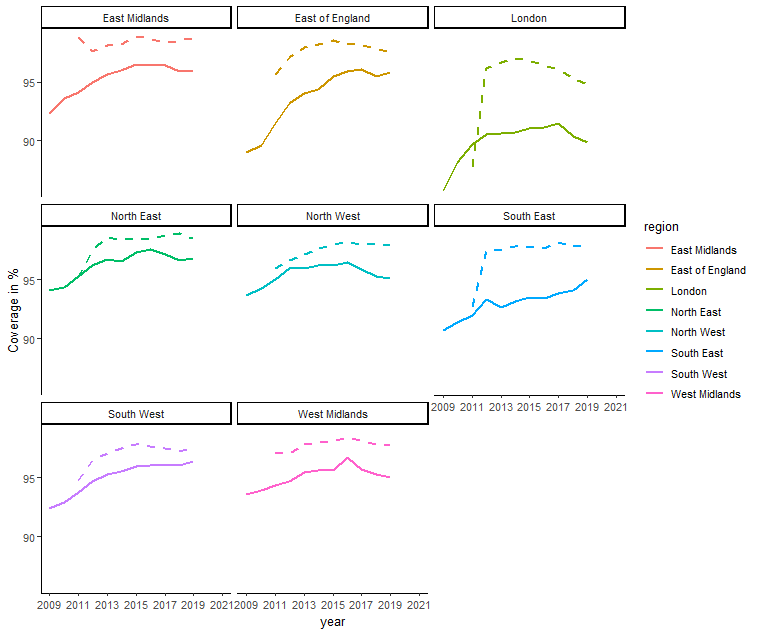

**Figure S2**. Comparing vaccine coverage by data source and region – MMR1 at the age of five for CPRD (dashed line) and COVER.

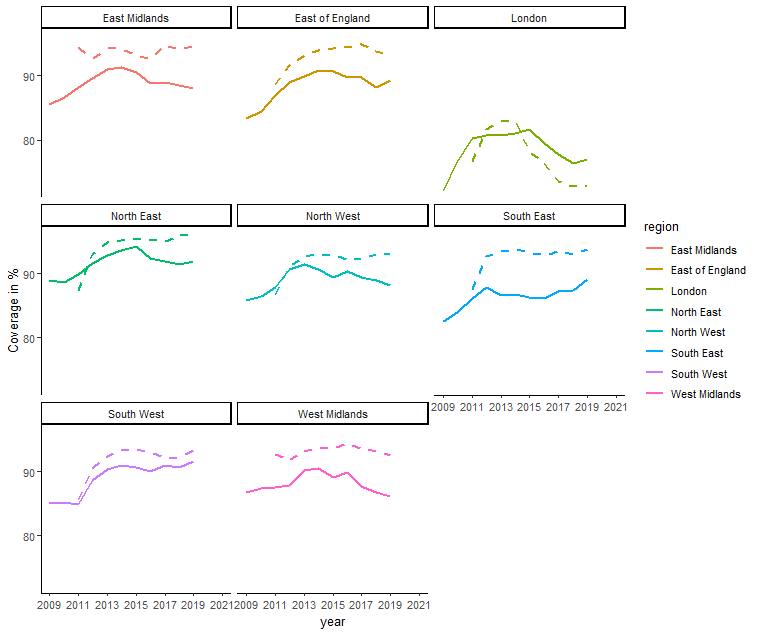
**Figure S3.** Comparing vaccine coverage by data source and region - – MMR2 at the age of five for CPRD (dashed line) and COVER.

**Table S4.** Comparison vaccine coverage at age five by data source

| **Year** | **MMR vaccine coverage in CPRD** | **National estimates for MMR in COVER** |
| --- | --- | --- |
| 2009-10 | - | 82.7 |
| 2010-11 | - | 84.2 |
| 2011-12 | 90.04 | 86.0 |
| 2012-13 | 90.78 | 87.7 |
| 2013-14 | 91.56 | 88.3 |
| 2014-15 | 91.23 | 88.6 |
| 2015-16 | 90.50 | 88.2 |
| 2016-17 | 89.91 | 87.6 |
| 2017-18 | 89.37 | 87.2 |
| 2018-19 | 89.20 | 86.4 |

**S5. Model description**

Overview

We used a deterministic, compartmental transmission model stratified by region and age group (<1;1-2;2-3;3-4;4-5;5-6;6-10;10-15;15-20;20-30;30-40;40+ years old), implemented with the R package odin.dust (23). In each age group and region, individuals were classified in several compartments: susceptible (i.e. not vaccinated nor previously infected), vaccinated once and protected, vaccinated once but failed to seroconvert, vaccinated twice and protected, vaccinated twice but failed to seroconvert, exposed (E), infectious (I) and recovered (i.e. previously infected). This is illustrated in Figure S6. On infection, the individuals moved to the exposed compartment, then they become infectious and finally recover. Depending on the vaccine coverage data, a proportion of individuals gained vaccination as they age. We assumed that vaccines provide protection according to an “all or nothing” principle (24,25): while a majority of newly vaccinated individuals gain full life-long protection, a proportion of individuals do not respond to the vaccine (primary vaccine failure). This proportion is estimated by the model.

Due to the short duration of exposure and infection in contrast to a year minimum spent in a compartment, there was no ageing in the exposed and infected compartments. The overall demographic structure was based on UK census data and simplified, ignoring migration between regions and countries and death in younger age groups.

We set the coverage of the first dose at the age of one to 75% of the coverage at age two. This was to represent that the first dose is usually given at the age of one, so children would have some protection between their first and second birthdays. Similarly, we set the coverage of the second dose at the age of three to 50% of the coverage at the age of four to account for the nine months of the fourth year of life which are spent vaccinated when the second dose is given at the age of three years and four months. This was to avoid bias in the model which would otherwise assume that there was no vaccination before age two and no second dose before age four due to the age bands in the model.

We assumed that no individuals in the 30-40 and 40+ age groups in 2010 was vaccinated, but the model estimated the proportion of individuals who gained immunity through infection.

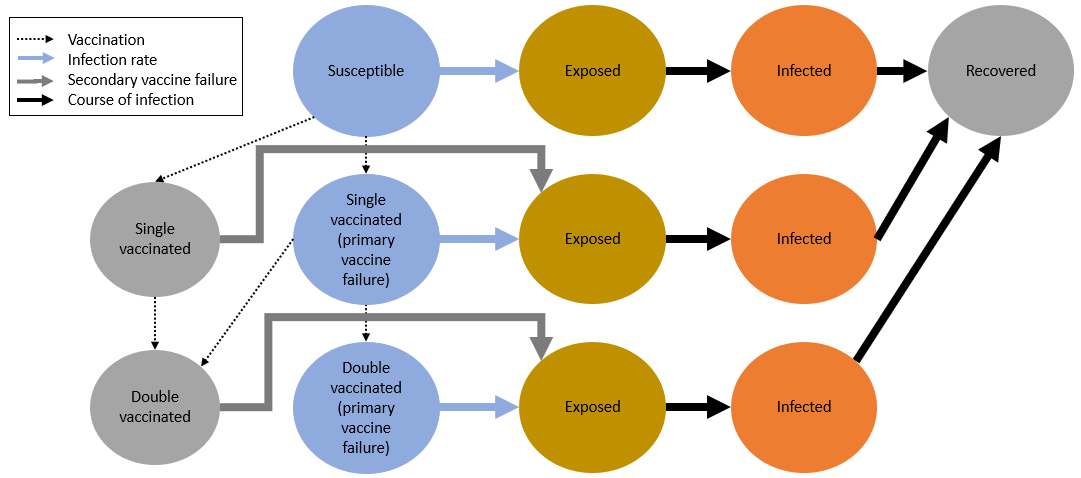

**Figure S6**. Structure of the model and its compartments. Reproduced from (Lancet citation).

**Table S7. Number of measles cases and cases avoided by vaccination strategy**

| **Vaccination Scenario** | **Median (IQR) number of cases across simulations** | **% of cases avoided in comparison to the median of the reference scenario (IQR)*** |
| --- | --- | --- |
| Reference | 7081 (6008 ;8388) | 0 (-18.46; 15.15) |
| *Scenarios of different vaccination schedules* | | |
| **MMR1 +0.25%** | 6523 (5524.25 ;7703.75) | 7.88 (-8.79; 21.98) |
| **MMR1 +0.5%** | 5921 (5066.5 ;6946.25) | 16.38 (1.90; 28.45) |
| **MMR1 +1%** | 5055.5 (4386.75 ;5871.5) | 28.60 (17.08; 38.05) |
| **MMR2 +1%** | 7085 (6009 ;8478) | -0.06 (-19.73; 15.14) |
| **MMR2 +3%** | 6889 (5771 ;8194) | 2.71 (-15.72; 18.5) |
| *Scenarios of different vaccination schedules* | | |
| **late MMR2** | 11868.5 (9541 ;14648.25) | -67.61 (-106.87; -34.74) |
| **early MMR2** | 5948 (5064 ;6944) | 16.00 (1.93; 28.48) |
| **early MMR2 with same speed of uptake as MMR1** | 5910 (5049.75 ;6929.5) | 16.54 (2.14; 28.69) |
| **early MMR2 with improved uptake +0.25%** | 5799 (4976 ;6812.25) | 18.10 (3.80; 29.73) |
| **Early MMR2 with improved uptake +0.5%** | 5740 (4909.75 ;6747.25) | 18.94 (4.71; 30.66) |
| **Early MMR2 with improved uptake +1%** | 5725.5 (4892.75 ;6785.25) | 19.14 (4.18; 30.9) |
| **Early MMR2 with coverage as MMR1** | 5402 (4611 ;6299.5) | 23.71 (11.04; 34.88) |
| **Early MMR2 with decreased uptake -3%** | 5952 (5083.5 ;7052.25) | 15.94 (0.41; 28.21) |
| **Early MMR2 with decreased uptake -5%** | 6169.5 (5240.5 ;7246.5) | 12.87 (-2.34; 25.99) |

*A positive percentage is the percentage of cases avoided; a negative percentage is the percentage of additional cases in comparison to the median of the reference scenario. The proportion of the cases avoided in comparison to the median was calculated as follows: Ncases-Median(reference)/Median(reference))*100.

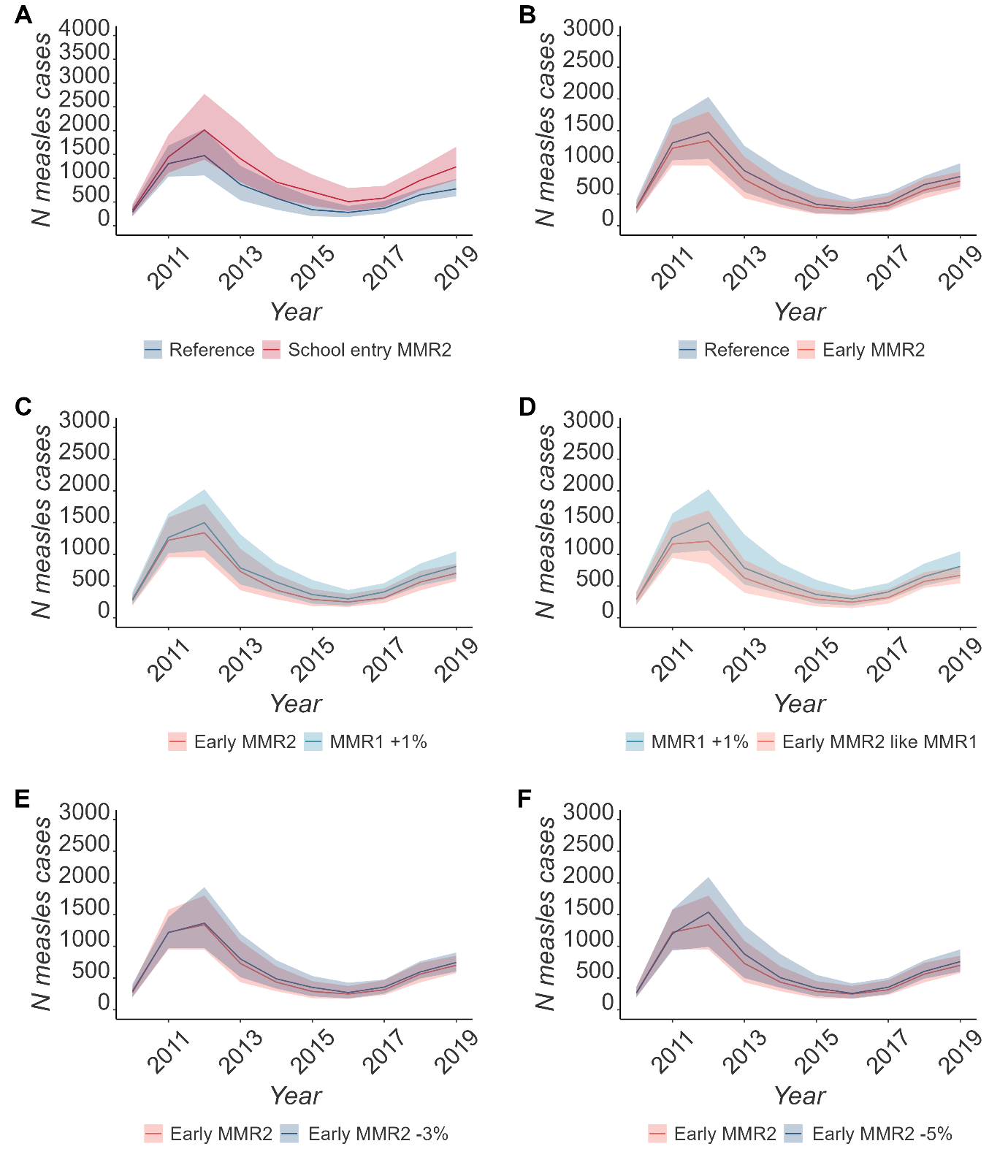

**Figure S8.** Comparing the median number and IQR of simulated cases across simulations using COVER data and including waning between (A) Reference scenario and MM2 given at school age, (B) Reference scenario and MMR2 given at the age of two, (C) MMR2 given at the age two against an increase of MMR1 by 1%., (D) increased MMR1 by 1% and an earlier MMR2 with the same coverage as MMR1. (E) and (F) are comparing the early MMR2 with the same uptake as before against a drop in coverage by 3% and 5% respectively.

**Table S9. Number of measles cases and cases avoided by vaccination strategy using COVER data instead of CPRD data**

| **Vaccination Scenario** | **Median (IQR) number of cases across simulations** | **% of cases avoided in comparison to the median of the reference scenario (IQR)*** |
| --- | --- | --- |
| Reference | 7305 (6321.5 ;8744.25) | 0 (-19.7; 13.46) |
| *Scenarios of different vaccination schedules* | | |
| **MMR1 +1%** | 7555.5 (6393 ;8891.5) | -3.43 (-21.72; 12.48) |
| *Scenarios of different vaccination schedules* | | |
| **late MMR2** | 10901 (9122 ;13192.5) | -49.23 (-80.6; -24.87) |
| **early MMR2** | 6599.5 (5568 ;7818.25) | 9.66 (-7.03; 23.78) |
| **Early MMR2 with improved uptake +1%** | 7050.5 (5969.25 ;8456) | 3.48 (-15.76; 18.29) |
| **Early MMR2 with coverage as MMR1** | 6326.5 (5411.75 ;7102.75) | 13.39 (2.77; 25.92) |
| **Early MMR2 with decreased uptake -3%** | 6925 (5912 ;8069.25) | 5.20 (-10.46; 19.07) |
| **Early MMR2 with decreased uptake -5%** | 7275 (5969 ;8649.75) | 0.41 (-18.41; 18.29) |

*A positive percentage is the percentage of cases avoided; a negative percentage is the percentage of additional cases in comparison to the median of the reference scenario. The proportion of the cases avoided in comparison to the median was calculated as follows: Ncases-Median(reference)/Median(reference))*100.

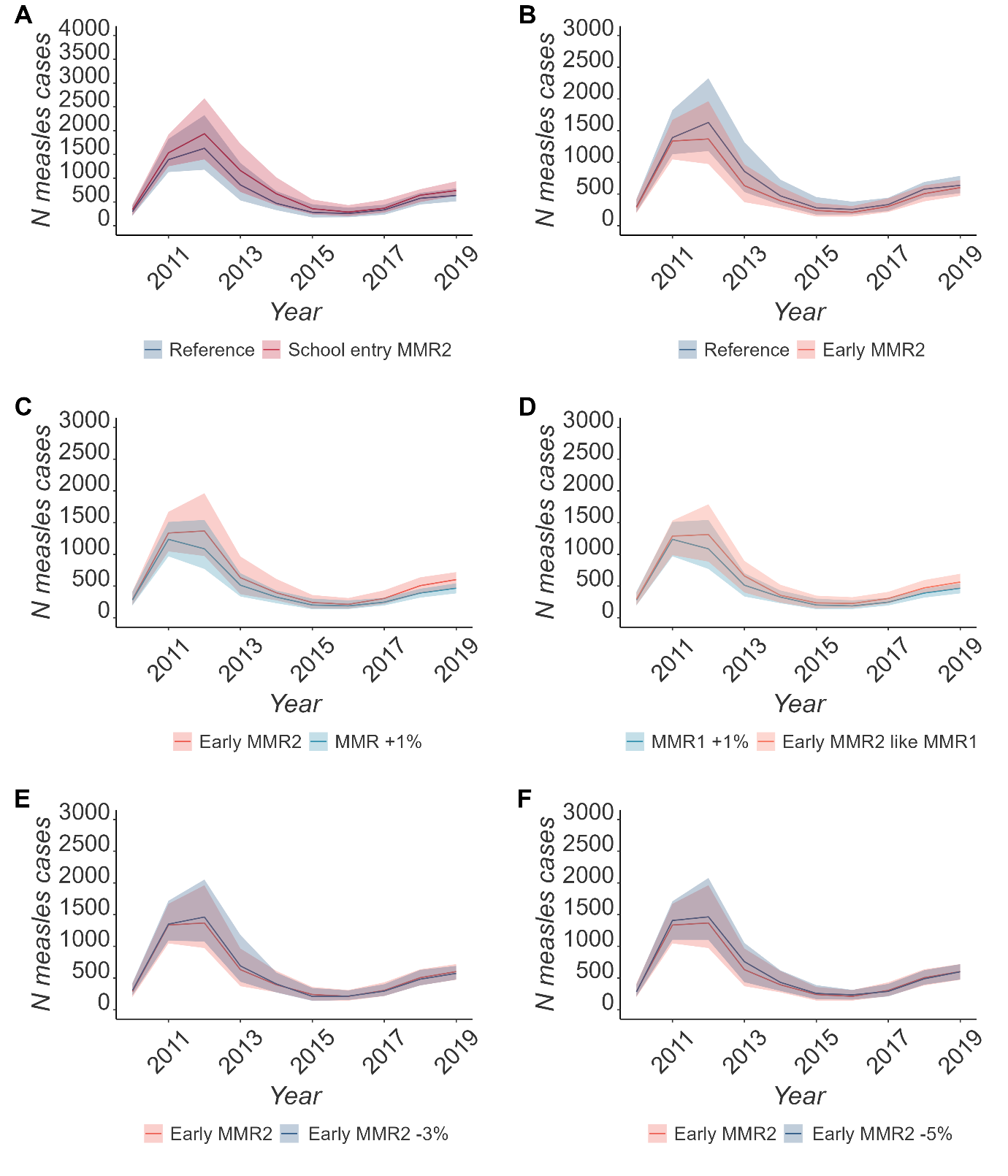

**Figure S10.** Comparing the median number and IQR of simulated cases across simulations using CPRD data and including waning between (A) Reference scenario and MM2 given at school age, (B) Reference scenario and MMR2 given at the age of two, (C) MMR2 given at the age two against an increase of MMR1 by 1%., (D) increased MMR1 by 1% and an earlier MMR2 with the same coverage as MMR1. (E) and (F) are comparing the early MMR2 with the same uptake as before against a drop in coverage by 3% and 5% respectively.

**Table S11**.Number of measles cases and cases avoided by vaccination strategy using CPRD data and including waning since vaccination into the final model.

| **Vaccination Scenario** | **Median (IQR) number of cases across simulations** | **% of cases avoided in comparison to the median of the reference scenario (IQR)*** |
| --- | --- | --- |
| Reference | 7157.5 (6001.25 ;8744) | 0 (-22.17; 16.15) |
| *Scenarios of different vaccination schedules* | | |
| **MMR1 +1%** | 5150 (4538.75 ;6007) | 28.05 (16.07; 36.59) |
| *Scenarios of different vaccination schedules* | | |
| **late MMR2** | 8694.5 (6993.5 ;10234) | -21.47 (-42.98; 2.29) |
| **early MMR2** | 6123.5 (5259.25 ;7263.75) | 14.45 (-1.48; 26.52) |
| **Early MMR2 with coverage as MMR1** | 5977 (5215.5 ;6949.25) | 16.49 (2.91; 27.13) |
| **Early MMR2 with decreased uptake -3%** | 6491 (5313.25 ;7687.5) | 9.31 (-7.42; 5.77) |
| **Early MMR2 with decreased uptake -5%** | 6434 (5646.75 ;7682.75) | 10.11 (-7.34; 21.11) |

*A positive percentage is the percentage of cases avoided; a negative percentage is the percentage of additional cases in comparison to the median of the reference scenario. The proportion of the cases avoided in comparison to the median was calculated as follows: Ncases-Median(reference)/Median(reference))*100.
